# Combining full-length gene assay and SpliceAI to interpret the splicing impact of all possible *SPINK1* coding variants

**DOI:** 10.1101/2023.11.14.23298498

**Authors:** Hao Wu, Jin-Huan Lin, Xin-Ying Tang, Wen-Bin Zou, Sacha Schutz, Emmanuelle Masson, Yann Fichou, Gerald Le Gac, Claude Férec, Zhuan Liao, Jian-Min Chen

**Affiliations:** Department of Gastroenterology, Changhai Hospital, Naval Medical University, Shanghai, China; Shanghai Institute of Pancreatic Diseases, Shanghai, China; Department of Prevention and Health Care, Eastern Hepatobiliary Surgery Hospital, Naval Medical University, Shanghai, China; Univ Brest, Inserm, EFS, UMR 1078, GGB, F-29200 Brest, France; CHRU Brest, F-29200 Brest, France

**Keywords:** Chronic pancreatitis, *In silico* prediction, Full-length gene splicing assay, Missense variant, Precision medicine, Pre-mRNA splicing, Single-nucleotide variant, SpliceAI, Splice site, *SPINK1*

## Abstract

**Background:** Single-nucleotide variants (SNVs) within gene coding sequences can significantly impact pre-mRNA splicing, bearing profound implications for pathogenic mechanisms and precision medicine. However, reliable splicing analysis often faces practical limitations, especially when the relevant tissues are challenging to access. While *in silico* predictions are valuable, they alone do not meet clinical classification standards. In this study, we aim to harness the well-established full-length gene splicing assay (FLGSA) in conjunction with SpliceAI to prospectively interpret the splicing effects of all potential coding SNVs within the four-exon *SPINK1* gene, a gene associated with chronic pancreatitis.

**Results:** We initiated the study with a retrospective correlation analysis (involving 27 previously FLGSA-analyzed *SPINK1* coding SNVs), progressed to a prospective correlation analysis (incorporating 35 newly FLGSA-tested *SPINK1* coding SNVs), followed by data extrapolation, and ended with further validation. In total, we analyzed 67 *SPINK1* coding SNVs, representing 9.3% of all 720 possible coding SNVs and affecting 19.2% of the 240 coding nucleotides. Among these 67 FLGSA-analyzed SNVs, 12 were found to impact splicing. Through extensive cross-correlation of the FLGSA-obtained and SpliceAI-predicted data, we reasonably extrapolated that none of the unanalyzed 653 coding SNVs in the *SPINK1* gene are likely to exert a significant effect on splicing. Out of these 12 splice-altering events, nine produced both wild-type and aberrant transcripts, while the remaining three exclusively generated aberrant transcripts. These splice-altering SNVs were predominantly concentrated in exons 1 and 2, particularly affecting the first and/or last coding nucleotide of each exon. Among the 12 splice-altering events, 11 were missense variants, constituting 2.17% of the 506 potential missense variants, while one was synonymous, accounting for 0.61% of the 164 potential synonymous variants.

**Conclusions:** Integrating FLGSA with SpliceAI, we conclude that less than 2% (1.67%) of all possible *SPINK1* coding SNVs have a discernible influence on splicing outcomes. Our findings underscore the importance of performing splicing analysis in the broader genomic sequence context of the study gene, highlight the inherent uncertainties associated with intermediate SpliceAI scores (i.e., those ranging from 0.20 to 0.80), and have general implications for the shift from “retrospective” to “prospective” analysis in terms of variant classification.

## Background

Single-nucleotide variants (SNVs) within the coding sequences of genes have the potential to exert a profound influence on pre-mRNA splicing. Remarkably, approximately 10% of disease-associated missense variants have been recognized as having the capacity to modulate pre-mRNA splicing [1]. This influence goes beyond missense variants and includes synonymous and nonsense variants [2, 3]. These findings have far-reaching implications for our understanding of disease pathogenesis and the advancement of precision medicine. For instance, what was once considered a ‘neutral’ missense variant or a ‘synonymous’ variant may, upon closer examination, be found to be disease-causing or related due to its impact on splicing. Similarly, the effectiveness of molecular treatment strategies targeting specific ‘missense’ or ‘nonsense’ variants may be compromised if these variants unexpectedly affect splicing.

The ideal approach for investigating the splicing effects of clinically detected SNVs is the analysis of RNA from pathophysiologically relevant tissues. However, practical constraints often limit access to these tissue samples [4]. As an alternative, RNA analysis from patient blood cells or immortalized lymphoblastoid cells is commonly employed, under the assumption that the gene of interest exhibits normal expression in these cell types [5]. When these options prove unfeasible, the frequently employed approach is the cell culture-based minigene splicing assay [6]. It is essential to acknowledge the inherent limitation of this assay – its inability to capture the broader genomic context of the study gene. This limitation could lead to erroneous findings [7, 8] due to the intricate nature of splicing regulation [9, 10].

In recent years, there have been significant advancements in the prediction of splicing outcomes for SNVs. An outstanding example is SpliceAI [11], a 32-layer deep neural network widely recognized as the most accurate tool for predicting splicing variants currently available (e.g., [12–15]). While these *in silico* prediction tools are valuable, they cannot be used in isolation to establish pathogenicity in accordance with variant classification guidelines recommended by the American College of Medical Genetics and Genomics (ACMG) [16]. Instead, they serve as first-line tools for variant classification and prioritization.

Another critical concern in the realm of precision medicine is the retrospective nature of functional analyses conducted on clinically identified variants [17]. This issue becomes increasingly urgent in an era when exome and genome sequencing have become integral to clinical diagnostics. Addressing this challenge requires a shift toward the prospective assessment of the functional impact of all potential SNVs at clinically significant loci in the human genome [18]. Multiplexed assays for variant effects (MAVE) offer a solution by enabling systematic collection of functional data for a multitude of variants in a single experiment [19]. A notable example is the prospective assessment of the functional impact, including splicing, of nearly 4,000 single nucleotide substitutions across 13 exons of the 24-exon *BRCA1* gene [20]. However, MAVE is technically and resource demanding, limiting its widespread application in many laboratories.

*SPINK1* (OMIM #167790) stands out as one of the primary genes associated with chronic pancreatitis [21]. Located on chromosome 5q32, the pathologically relevant *SPINK1* mRNA isoform (NM_001379610.1) comprises four exons, encoding a 79-amino acid precursor protein that eventually yields the mature 56-amino-acid pancreatic secretory trypsin [22, 23]. Loss-of-function variants in the *SPINK1* gene increase susceptibility to chronic pancreatitis through the trypsin-dependent pathway [21, 24, 25]. Previously, we successfully cloned the ∼7-kb genomic sequence of the four-exon *SPINK1* gene into the pcDNA3.1/V5-His-TOPO vector, establishing a cell culture-based full-length gene splicing assay (FLGSA) [26]. Notably, FLGSA, unlike the frequently used minigene assay, preserves the broader natural genomic context of the gene under investigation—a crucial factor considering the intricacies of splicing regulation. Naturally, FLGSA also provides a practical advantage over the minigene assay, enabling comprehensive analysis of all coding and intronic variants within a consistent genomic framework.

In the context of the *SPINK1* gene, we have previously employed the FLGSA assay to analyze both known coding and intronic variants [7, 27–31]. The accuracy of the FLGSA assay is illuminated by the study of the *SPINK1* c.194+2T>C variant, a type of variant often considered to cause a complete functional loss of the affected allele due to its occurrence within the canonical GT splice donor site [32]. Specifically, the findings from the FLGSA assay [27] were in alignment with *in vivo* splicing data [33] for c.194+2T>C, revealing a notable presence of wild-type (WT) transcripts alongside with exon 3-skipping aberrant transcripts (N.B. the ratio of WT transcripts to aberrant transcripts was subsequently estimated to be 1:9 [34]). Remarkably, this preservation of 10% residual function was associated with the less severe phenotypes observed in *SPINK1* c.194+2T>C homozygotes, who exhibit chronic pancreatitis with variable expressivity [35]. In contrast, homozygous *SPINK1* variants leading to a complete 100% loss of the gene product are linked to a more severe phenotype referred to as severe infantile isolated exocrine pancreatic insufficiency [36].

In this study, we set out to harness the combined power of the FLGSA assay and SpliceAI’s predictive capabilities to prospectively interpret the splicing effects of all potential coding SNVs within the *SPINK1* gene.

## Methods

### Research rationale and strategy

The primary objective of this study was to prospectively interpret the splicing impact of all potential coding SNVs within the *SPINK1* gene by leveraging a synergistic combination of the FLGSA assay and SpliceAI predictions. Our hypothesis was grounded in the belief that insights derived from correlating experimental data obtained through FLGSA with SpliceAI predictions for a subset of *SPINK1* coding SNVs could be reasonably extrapolated to the broader pool of unanalyzed *SPINK1* coding SNVs. The study would begin with a retrospective correlation analysis (using previously FLGSA-analyzed *SPINK1* coding SNVs), advance to a prospective correlation analysis (involving newly FLGSA-tested *SPINK1* coding SNVs), followed by data extrapolation, and end with further validation (Fig. 1).

**Figure 1.**
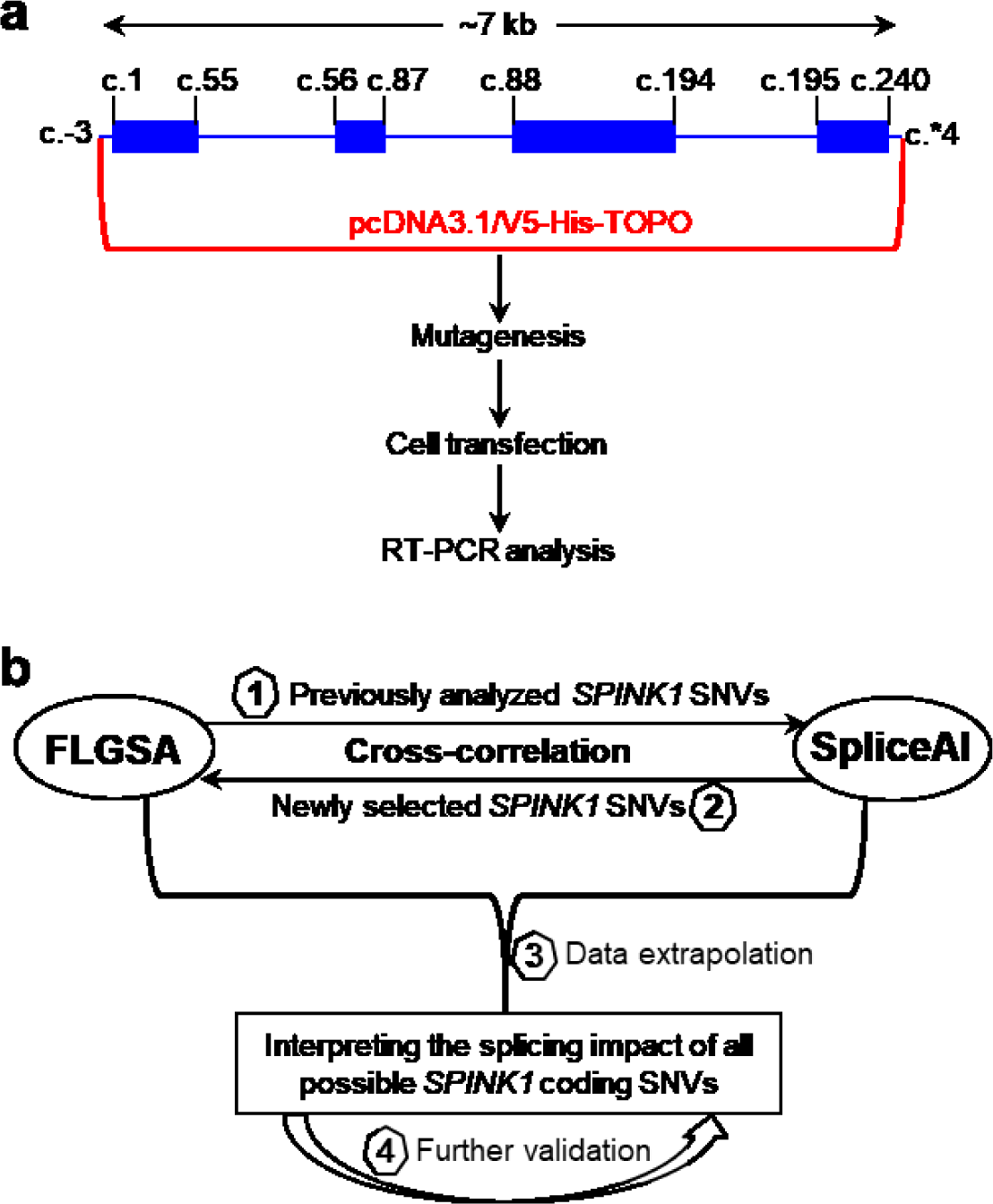
Overview of the FLGSA assay and research strategy. **a** Representation of the *SPINK1* full-length gene expression vector and the experimental steps involved in the FLGSA assay for each study variant. The coding sequences of the four-exon *SPINK1* gene are to scale, while the intronic and untranslated region sequences are not. The reference *SPINK1* genomic sequence is NG_008356.2, and the reference *SPINK1* mRNA sequence is MANE (Matched Annotation from the NCBI and EMBL-EBI [37]) select ENST00000296695 or NM_001379610.1. NM_001379610.1 represents the *SPINK1* transcript isoform expressed in the exocrine pancreas. The starting and ending positions of the coding sequences in each exon, as well as those of the *SPINK1* genomic sequence cloned into the pcDNA3.1/V5-His-TOPO vector, are indicated in accordance with NM_001379610. **b** Illustration demonstrating how the FLGSA assay was integrated with SpliceAI to prospectively evaluate the splicing effects of all potential coding variants within the *SPINK1* gene. Abbreviations: FLGSA, full-length gene splicing assay; RT-PCR, reverse transcription-PCR; SNVs, single-nucleotide variants.

### SpliceAI

SpliceAI provides four Δ scores: acceptor gain (AG), acceptor loss (AL), donor gain (DG), and donor loss (DL). These scores represent the maximum difference between the probability of the variant and the reference alleles concerning splice-altering. The Δ score ranges from 0 to 1, with higher scores indicating a greater likelihood that the variant affects splicing. Variants with a Δ score of <0.20 were generally considered unlikely to have a substantial impact on splicing, while variants with a Δ score exceeding 0.80 were generally associated with a high specificity for splicing alterations [11]. SpliceAI also provides the pre-mRNA positions of the predicted splicing effect with respect to the variant position. For *SPINK1* variants, positive and negative pre-mRNA positions indicate positions 5’ and 3’ to the variant position in terms of the gene’s sense strand.

Our retrospective analysis involved comparing FLGSA data with SpliceAI predictions for known *SPINK1* coding SNVs. For our prospective analysis, we selected new *SPINK1* coding SNVs for FLGSA analysis based on SpliceAI-predicted Δ scores. These steps relied on SpliceAI Δ scores obtained from [38] using the default settings of SpliceAI in February 2020. These SpliceAI Δ scores correspond to Illumina precomputed scores created using Gencode v24 and max distance = 50bp at the time [11] and align with those accessible to academic users on the SpliceAI Virtual website [39]. Importantly, in May 2023, SpliceAI retired these Illumina precomputed scores. To adapt to this change and refine the cross-correlation, we additionally conducted a second-step analysis for *SPINK1* coding SNVs that underwent the FLGSA assay, utilizing SpliceAI Δ scores obtained from SpliceAI Lookup [40] with the following parameters: (i) Genome version, hg38; (ii) Score type, Raw; and (iii) Max distance, 10,000. This new set of SpliceAI Δ scores was manually obtained in October 2023.

### Collation of known *SPINK1* coding variants with FLGSA data

To date, the FLGSA assay has been employed to analyze 27 clinically identified *SPINK1* coding SNVs, comprising 24 missense variants and 3 synonymous variants [7, 31]. All these 27 variants were included in our retrospective correlation analysis.

### Selection of potential *SPINK1* coding variants for FLGSA

We conducted a rigorous selection process to identify potential *SPINK1* coding variants for FLGSA. This process involved a comprehensive assessment of Illumina precomputed SpliceAI Δ scores for all 720 potential coding SNVs, which arose from the multiplication of 240 coding nucleotides by 3, within the *SPINK1* gene. As a general guideline, we chose to encompass all three possible SNVs at the beginning (with the exception of exon 1) and end (excluding exon 4) of each exon, regardless of their SpliceAI Δ scores. Additionally, we included SNVs with at least one SpliceAI Δ score ≥0.20, excluding those deemed physiologically irrelevant. Furthermore, we incorporated some variants predicted to have no impact on splicing as controls. This process initially led us to select 35 SNVs. For the purpose of further validation, we selected additional five SNVs for FLGSA. More detailed information is provided in the *Results* section.

### FLGSA

The newly selected *SPINK1* coding SNVs underwent FLGSA analysis, as previously described [27, 29, 32]. Specifically, the introduction of the selected variants into the full-length gene expression vector containing the WT *SPINK1* genomic sequence [26] and the subsequent confirmation of the introduced variants through Sanger sequencing were executed by GENEWIZ Biotech Co. (Suzhou, China). All subsequent experimental procedures were conducted at the Shanghai Changhai laboratory.

#### Cell culture, transfection, RNA extraction, and reverse transcription (RT)

Human embryonic kidney 293T (HEK293T) cells were cultured in the DMEM basic medium (Gibco) with 10% fetal calf serum (Procell). 3.5 × 10^5^ cells were seeded per well in 6-well plates 24 hours before transfection. 2.5 µg of either WT or variant plasmid, mixed with HieffTrans Universal Transfection Reagent (Yeasen), was used for transfection per well. Forty-eight hours after transfection, total RNA was extracted using the FastPure Cell/Tissue Total RNA Isolation Kit V2 (+gDNA wiper) (Vazyme). RT was carried out using the HiScript III 1st Strand cDNA Synthesis Kit (Vazyme), incorporating 2 µL of 5 × gDNA wiper Mix, 2 µL of 10 × RT Mix, 2 µL of HiScript III Enzyme Mix, 1 µL of Oligo (dT)20VN, and 1 µg of total RNA.

#### RT-PCR and sequencing of the resulting products

RT-PCR was performed in a 25-μL reaction mixture containing 12.5 μL 2 × Taq Master Mix (Vazyme), 1 μL cDNA, and 0.4 μM of each primer. The primers used were 5’-GGAGACCCAAGCTGGCTAGT-3’ (forward) and 5’-AGACCGAGGAGAGGGTTAGG-3’ (reverse), both of which are located within the pcDNA3.1/V5-His-TOPO vector sequence. The PCR program had an initial denaturation step at 94°C for 5 min, followed by 35 cycles of denaturation at 94°C for 30 s, annealing at 55°C for 30 s, and extension at 72°C for 5 min, and a final extension step at 72°C for 7 min. RT-PCR products presenting either a single band or multiple bands were excised from the agarose gel and then purified using a Gel Extraction Kit (Omega Bio-Tek). The sequencing primers employed were identical to those used for the RT-PCR analyses. Sequencing reactions were conducted using the BigDye Terminator v3.1 Cycle Sequencing Kit (Applied Biosystems).

#### Approximate estimate of relative expression levels of co-expressed WT and aberrant transcripts

To estimate the relative expression levels of aberrantly spliced transcripts in comparison to WT transcripts for variants that produced both types of transcripts, we utilized ImageJ [41].

### The contribution of generative artificial intelligence to the writing process

We used ChatGPT-4 [42] to enhance the readability and linguistic quality of this manuscript. We take full responsibility for the content presented herein.

## Results

### Retrospective correlation of FLGSA data with SpliceAI predictions for known *SPINK1* coding SNVs

We initiated the study with a retrospective analysis involving known *SPINK1* coding SNVs that had previously undergone FLGSA analysis. All 27 such variants consistently yielded WT transcripts in the FLGSA assay [7, 31]. Details of these variants, including their precomputed SpliceAI Δ scores by Illumina [11], are provided in Table 1 (see the end of the manuscript).

**Table 1.** FLGSA data and Illumina precomputed SpliceAI Δ scores for 27 known and 35 potential *SPINK1* coding SNVs.

| Exon | Variant<sup>a</sup> |  | Illumina precomputed SpliceAI scores<sup>b</sup> |  |  |  | Generation of aberrant transcripts as determined by FLGSA<sup>c</sup> | Study |
| --- | --- | --- | --- | --- | --- | --- | --- | --- |
|  | Nucleotide change | Amino acid change | AG | AL | DG | DL |  |  |
| 1 | c.9A>C | p.Val3Val | 0.00 | 0.00 | 0.04 | 0.55 (3 bp)<sup>d</sup> | No | This study |
| 1 | c.9A>G | p.Val3Val | 0.00 | 0.00 | 0.04 | 0.54 (3 bp)<sup>d</sup> | No | This study |
| 1 | c.9A>T | p.Val3Val | 0.00 | 0.00 | 0.04 | 0.55 (3 bp)<sup>d</sup> | No | This study |
| 1 | c.11C>G | p.Thr4Arg | 0.00 | 0.00 | 0.44 (5 bp) | 0.45 (-44 bp) | No | This study |
| 1 | c.15C>T | p.Gly5Gly | 0.00 | 0.00 | 0.52 (2 bp) | 0.13 (-40 bp) | No | This study |
| 1 | c.26T>G | p.Leu9Arg | 0.00 | 0.00 | 0.03 | 0.29 (20 bp)<sup>d</sup> | No | [31] |
| 1 | c.29G>A | p.Ser10Asn | 0.00 | 0.00 | 0.11 (23 bp) | 0.03 | No | [31] |
| 1 | c.36G>C | p.Leu12Phe | 0.00 | 0.00 | 0.02 | 0.10 (30 bp)<sup>d</sup> | No | [31] |
| 1 | c.41T>C | p.Leu14Pro | 0.00 | 0.00 | 0.01 | 0.04 | No | [31] |
| 1 | c.41T>G | p.Leu14Arg | 0.00 | 0.00 | 0.02 | 0.09 (35 bp)<sup>d</sup> | No | [31] |
| 1 | c.43T>A | p.Leu15Met | 0.00 | 0.00 | 0.02 | 0.12 (37 bp)<sup>d</sup> | No | This study |
| 1 | c.43T>C | p.Leu15= | 0.00 | 0.00 | 0.03 | 0.14 (37 bp)<sup>d</sup> | No | This study |
| 1 | c.43T>G | p.Leu15Val | 0.00 | 0.00 | 0.38 (1 bp) | 0.00 | No | This study |
| 1 | c.55G>A | p.Gly19Ser | 0.00 | 0.00 | 0.36 (49 bp) | 0.40 (0 bp) | Yes (Intron 1 retention<sup>e</sup>/WT: 1/9.03) | This study |
| 1 | c.55G>C | p.Gly19Arg | 0.00 | 0.00 | 0.33 (49 bp) | 0.34 (0 bp) | Yes (Intron 1 retention<sup>e</sup>/WT: 1/21.72) | This study |
| 1 | c.55G>T | p.Gly19Cys | 0.00 | 0.00 | 0.37 (49 bp) | 0.51 (0 bp) | Yes (Intron 1 retention<sup>e</sup>/WT: 1/9.38) | This study |
| 2 | c.56G>A | p.Gly19Asp | 0.01 | 0.10 (0 bp) | 0.00 | 0.07 | No | This study |
| 2 | c.56G>C | p.Gly19Ala | 0.01 | 0.40 (0 bp) | 0.00 | 0.29 (-31 bp) | Yes (E2 skipping/WT: 1/1.32) | This study |
| 2 | c.56G>T | p.Gly19Val | 0.01 | 0.61 (0 bp) | 0.00 | 0.46 (-31 bp) | Yes (E2 skipping/WT: 2.97/1) | This study |
| 2 | c.65G>T | p.Gly22Val | 0.00 | 0.31 (9 bp) | 0.00 | 0.17 (-22 bp) | Yes (E2 skipping/WT: 1/5.16) | This study |
| 2 | c.75C>T | p.Ser25= | 0.00 | 0.02 | 0.00 | 0.02 | No | [31] |
| 2 | c.80G>T | p.Gly27Val | 0.00 | 0.09 | 0.61 (2 bp) | 0.10 (-7 bp) | No | This study |
| 2 | c.84A>C | p.Arg28Ser | 0.00 | 0.01 | 0.00 | 0.00 | No | This study |
| 2 | c.84A>G | p.Arg28= | 0.00 | 0.49 (28 bp) | 0.00 | 0.25 (-3 bp) | Yes (E2 skipping/WT: 10.80/1) | This study |
| 2 | c.84A>T | p.Arg28Ser | 0.00 | 0.04 | 0.01 | 0.02 | No | This study |
| 2 | c.85G>T | p.Glu29* | 0.00 | 0.25 (29 bp) | 0.00 | 0.17 (-2 bp) | No | This study |
| 2 | c.86A>C | p.Glu29Ala | 0.00 | 0.51 (30 bp) | 0.01 | 0.23 (-1 bp) | No | This study |
| 2 | c.86A>G | p.Glu29Gly | 0.00 | 0.84 (30 bp) | 0.00 | 0.67 (-1 bp) | Yes (E2 skipping/WT: 4.13/1) | This study |
| 2 | c.86A>T | p.Glu29Val | 0.00 | 0.81 (30 bp) | 0.00 | 0.58 (-1 bp) | Yes (E2 skipping/WT: 1/5.31) | This study |
| 2 | c.87G>A | p.Glu29= | 0.00 | 0.87 (31 bp) | 0.00 | 0.93 (0 bp) | Yes (Complete E2 skipping) | This study |
| 2 | c.87G>C | p.Glu29Asp | 0.00 | 0.84 (31 bp) | 0.01 | 0.93 (0 bp) | Yes (Complete E2 skipping) | This study |
| 2 | c.87G>T | p.Glu29Asp | 0.00 | 0.88 (31 bp) | 0.02 | 0.93 (0 bp) | Yes (Complete E2 skipping) | This study |
| 3 | c.88G>A | p.Ala30Thr | 0.00 | 0.00 | 0.00 | 0.00 | No | This study |
| 3 | c.88G>C | p.Ala30Pro | 0.00 | 0.00 | 0.00 | 0.00 | No | This study |
| 3 | c.88G>T | p.Ala30Ser | 0.00 | 0.01 | 0.00 | 0.00 | No | This study |
| 3 | c.101A>G | p.Asn34Ser | 0.00 | 0.00 | 0.00 | 0.00 | No | [31] |
| 3 | c.110A>G | p.Asn37Ser | 0.00 | 0.00 | 0.00 | 0.00 | No | [31] |
| 3 | c.123G>C | p.Lys41Asn | 0.00 | 0.00 | 0.00 | 0.00 | No | [31] |
| 3 | c.126A>G | p.Ile42Met | 0.00 | 0.00 | 0.00 | 0.00 | No | [31] |
| 3 | c.133C>T | p.Pro45Ser | 0.00 | 0.00 | 0.00 | 0.00 | No | [31] |
| 3 | c.137T>A | p.Val46Asp | 0.00 | 0.00 | 0.00 | 0.00 | No | [31] |
| 3 | c.143G>A | p.Gly48Glu | 0.00 | 0.00 | 0.00 | 0.00 | No | [31] |
| 3 | c.150T>G | p.Asp50Glu | 0.00 | 0.00 | 0.00 | 0.00 | No | [31] |
| 3 | c.160T>C | p.Tyr54His | 0.00 | 0.00 | 0.00 | 0.00 | No | [31] |
| 3 | c.163C>T | p.Pro55Ser | 0.00 | 0.00 | 0.00 | 0.00 | No | [31] |
| 3 | c.174C>T | p.Cys58= | 0.00 | 0.00 | 0.00 | 0.00 | No | [31] |
| 3 | c.178T>G | p.Leu60Val | 0.00 | 0.00 | 0.26 (1 bp) | 0.00 | No | This study |
| 3 | c.190A>G | p.Asn64Asp | 0.00 | 0.00 | 0.00 | 0.00 | No | [31] |
| 3 | c.193C>T | p.Arg65Trp | 0.00 | 0.00 | 0.00 | 0.00 | No | [31] |
| 3 | c.194G>A | p.Arg65Gln | 0.00 | 0.00 | 0.00 | 0.00 | No | [7] |
| 3 | c.194G>C | p.Arg65Pro | 0.00 | 0.00 | 0.00 | 0.00 | No | This study |
| 3 | c.194G>T | p.Arg65Leu | 0.00 | 0.00 | 0.00 | 0.00 | No | This study |
| 4 | c.195G>A | p.Arg65= | 0.00 | 0.03 | 0.00 | 0.00 | No | This study |
| 4 | c.195G>C | p.Arg65= | 0.00 | 0.10 (0 bp) | 0.00 | 0.00 | No | This study |
| 4 | c.195G>T | p.Arg65= | 0.00 | 0.23 (0 bp) | 0.00 | 0.00 | No | This study |
| 4 | c.198A>C | p.Lys66Asn | 0.01 | 0.00 | 0.00 | 0.00 | No | [31] |
| 4 | c.199C>T | p.Arg67Cys | 0.00 | 0.01 | 0.00 | 0.00 | No | [31] |
| 4 | c.200G>A | p.Arg67His | 0.00 | 0.00 | 0.00 | 0.00 | No | [31] |
| 4 | c.203A>G | p.Gln68Arg | 0.01 | 0.00 | 0.00 | 0.00 | No | [31] |
| 4 | c.206C>T | p.Thr69Ile | 0.00 | 0.01 | 0.00 | 0.00 | No | [31] |
| 4 | c.231G>A | p.Gly77= | 0.01 | 0.00 | 0.00 | 0.00 | No | [31] |
| 4 | c.236G>T | p.Cys79Phe | 0.00 | 0.02 | 0.00 | 0.00 | No | [31] |

*Abbreviations:* AG, acceptor gain; AL, acceptor loss; DG, donor gain; DL, donor loss; FLGSA, full-length gene splicing assay; SNVs, single-nucleotide variants; WT, wild-type
<sup>a</sup>*SPINK1* mRNA reference sequence: NM\_001379610.1.
<sup>b</sup>In parentheses: corresponding pre-mRNA positions are provided for $\Delta$ scores $\geq 0.10$ . Positive and negative pre-mRNA positions indicate positions 5' and 3' to the variant position in terms of the gene's sense strand.
<sup>c</sup>In parentheses: in case of the generation of aberrant transcripts, the nature of the aberrant transcripts and the ratio of aberrant transcripts to WT transcripts are indicated.
<sup>d</sup>Considered not to be physiologically relevant as the predicted donor loss is situated within exon 1 of the *SPINK1* gene.
<sup>e</sup>Retention of the first 140 bases of intron 1.

Among the 108 corresponding SpliceAI Δ scores, only one exceeded the threshold of 0.20, specifically, a DL Δ score of 0.29 (20 bp) for c.26T>G. It’s important to note that this DL score was not physiologically relevant, as the predicted donor loss pertains to the GT dinucleotide at *SPINK1* coding positions c.7_8. Additionally, it’s worth mentioning that none of the four alternative *SPINK1* transcript isoforms (NM_003122.5, NM_001354966.2, XM_047417625.1, and XM_047417626.1 [43]) utilizes the c.7_8 GT dinucleotide as a splice donor site. The next highest score was a mere 0.11, specifically a DG score for c.29G>A. Therefore, except for the case of c.26T>G, a perfect correlation was observed between the FLGSA-derived and SpliceAI-predicted data in the context of the subset of known *SPINK1* coding SNVs.

### Selection of potential *SPINK1* coding SNVs for FLGSA

Next, we aimed to select a new set of *SPINK1* coding SNVs for FLGSA analysis. We based our selection on the Illumina precomputed SpliceAI scores (Supplementary Table S1). Adhering to the methodological guidelines detailed in the *Methods* section, we carefully chose 35 SNVs. To enhance clarity, we will elucidate the selection process within the context of the four exons. Details of the 35 chosen SNVs, alongside their corresponding Illumina precomputed SpliceAI scores, are provided in Table 1.

Exon 1, comprising 55 coding nucleotides, exhibited AG and AL scores of zero for all 165 possible SNVs. As a result, our selection process relied on DG and DL scores. Initially, we included all three possible SNVs at the terminal position of exon 1 (i.e., c.55G>A, c.55G>C, and c.55G>T), whose DG and DL scores ranged from 0.33 to 0.51. Subsequently, from the remaining SNVs, we included the three with the highest DG scores (i.e., c.11C>G, 0.44; c.15C>T, 0.52; and c.43T>G, 0.38), along with two additional variants at c.43, specifically c.43T>C and c.43T>A. Regarding DL scores, those with positive values (referring to positions within either the 5’-UTR or coding sequence of exon 1) and some with negative values (referring to positions within the coding sequence of exon 1) were deemed physiologically irrelevant. Notably, all five variants with a physiologically relevant DL score of >0.10 (i.e., c.11C>G, 0.45; c.15C>T, 0.13; c.55G>A, 0.40; c.55G>C, 0.34; and c.55G>T, 0.51) also possessed a DG score of at least 0.33, and hence, were already included in our analysis. Lastly, for the purpose of comparison, we included all three possible SNVs at c.9, all of which were predicted to have a DG score of 0.04.

Exon 2, encompassing 32 coding nucleotides, hosts 96 potential SNVs. Notably, two SNVs at the starting position (c.56) and all three SNVs at the final two positions (c.86 and c.87) of exon 2 demonstrated AL and DL scores >0.20. As a result, we included all potential SNVs at these three positions in the functional analysis. Additionally, the sole additional SNV meeting the criteria of both AL and DL scores >0.20 was c.84A>G. Hence, we incorporated this SNV, along with the other two possible SNVs at the c.84 position, into the functional analysis. Finally, four variants demonstrated a single score surpassing 0.20. These included c.64G>T with an AL score of 0.22, c.65G>T with an AL score of 0.31, c.80G>T with a DG score of 0.61, and c.85G>T with an AL score of 0.25. For the FLGSA assay, we selected the latter three variants for inclusion.

Exon 3, comprising 107 coding nucleotides, contains 321 potential SNVs. Among the 1284 SpliceAI scores associated with these variants, most were zero. However, exceptions included an AG score of 0.13 for c.92A>G and a DG score of 0.26 for c.178T>G. In our FLGSA analysis, we prioritized c.178T>G. Additionally, we incorporated three SNVs at the beginning of exon 3 (c.88G>A, c.88G>C, and c.88G>T) and two at its end (c.194G>C and c.194G>T, with the note that c.194G>A had been previously analyzed in [7]).

Exon 4, with 46 coding nucleotides, contains 138 possible SNVs. All 552 corresponding SpliceAI Δ scores consistently remained at or near zero, with a maximum of 0.05, except for two cases: an AL score of 0.10 for c.195G>C and an AL score of 0.23 for c.195G>T. Since c.195 is the starting position of exon 4, we included all three possible SNVs at this position for FLGSA.

### FLGSA assay for the 35 prospectively selected *SPINK1* coding SNVs

Then, we embarked on the functional characterization of the splicing impact of the aforementioned 35 prospective *SPINK1* coding SNVs using the FLGSA assay. The outcomes, represented by RT-PCR band patterns in agarose gel analysis, are detailed in Fig. 2. In accordance with our common practice [27, 29, 30], we employed Sanger sequencing to determine the identity of RT-PCR bands whenever possible. This step carried particular significance for two primary reasons: (i) a seemingly WT RT-PCR band could differ from the genuine WT by only one or two base pairs [30, 32], and (ii) this information was pivotal for comparison with SpliceAI-predicted splice-altering sites in instances of aberrant splicing. Additionally, it’s worth mentioning that the WT transcripts originating from cells transfected with the variant expression vectors consistently contained the corresponding coding SNVs.

**Figure 2.**
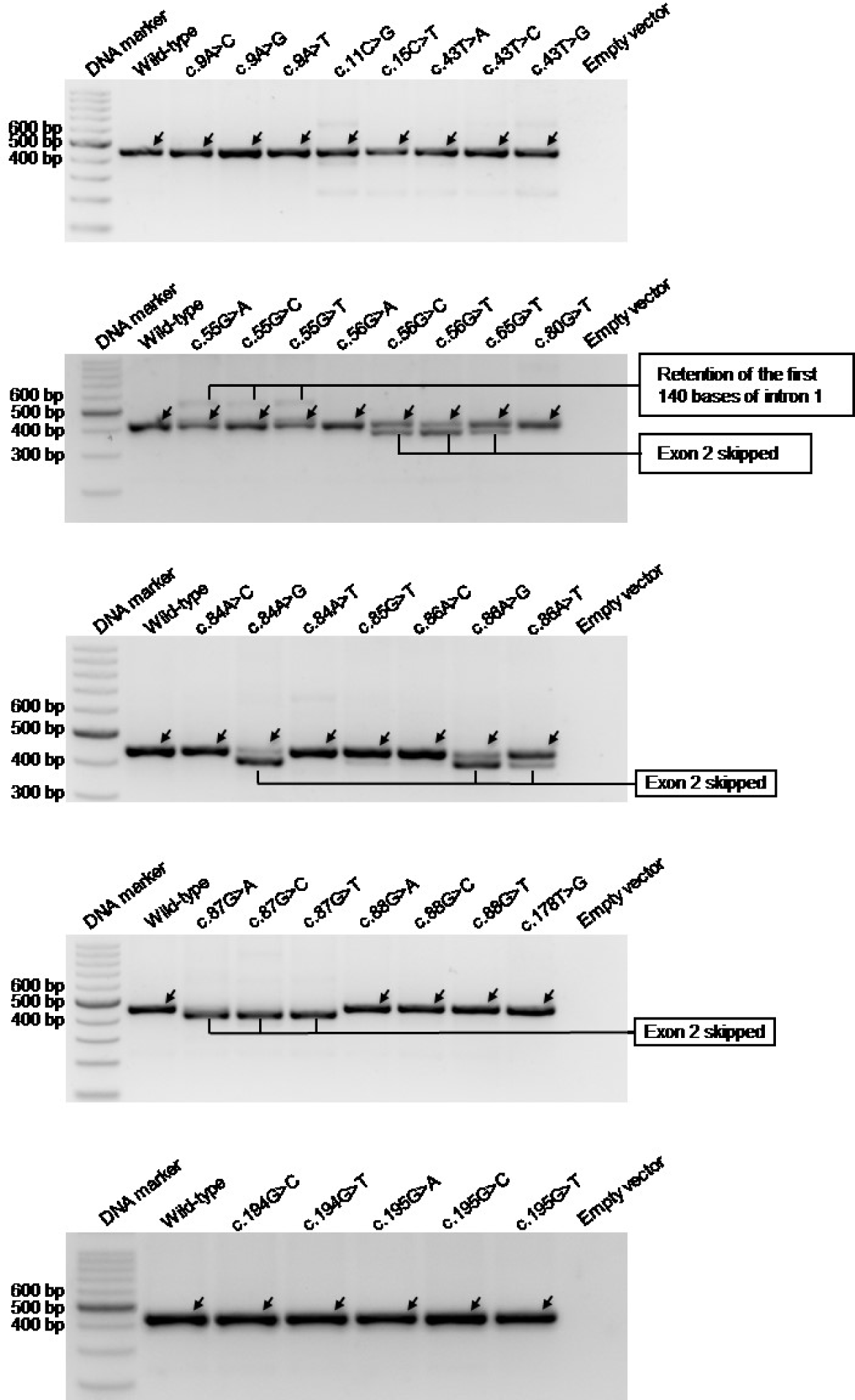
RT-PCR results for the FLGSA analysis of 35 potential *SPINK1* coding variants. In all panels, arrows indicate wild-type transcripts, all of which were confirmed by Sanger sequencing. FLGSA, full-length gene splicing assay. RT-PCR, reverse transcription-PCR.

However, we encountered challenges in sequencing several faint bands (e.g., the barely discernible band below the major band in c.11C>G; Fig. 2). These faint bands could potentially signify authentic aberrantly spliced transcripts or spurious amplifications. It is important to note two key considerations. Firstly, our prior investigation of 5’ splice site GT>GC variants established that our FLGSA assay was capable of detecting as little as ∼1% of normally spliced transcripts [32]. Secondly, we had previously postulated that a *SPINK1* variant is unlikely to have pathological relevance if it caused a <10% loss of normal function (or retained >90% normal function) [25]. Therefore, we excluded these barely visible RT-PCR bands from further consideration.

A concise summary of the FLGSA findings, including comparative levels of aberrant versus WT transcripts where applicable, can be found in Table 1.

### Correlation of FLGSA data with SpliceAI predictions for the 35 prospectively analyzed SNVs

Subsequently, we conducted a thorough analysis to correlate the FLGSA data generated for the prospectively examined 35 SNVs with their corresponding Illumina precomputed SpliceAI scores (Table 1). After discarding physiologically irrelevant DL scores linked to various SNVs in exon 1, we observed a significant pattern in relation to the threshold Δ scores of 0.20 and 0.80 [11]. Specifically, all variants with a Δ score not exceeding 0.20 exclusively produced WT transcripts. Conversely, every variant with a Δ score above 0.80 consistently led to the production of aberrant variants.

Establishing a clear correlation between the presence or absence of aberrant transcripts and intermediate Δ scores (0.20 to 0.80) was challenging. However, a notable pattern emerged upon analyzing Δ scores for SNVs at positions c.55, c.56, and c.86. Each of these positions underwent FLGSA analysis, with at least two of the three possible SNVs at each position generating both aberrant and WT transcripts. For a more focused analysis, we will compare the DL scores associated with these SNVs. At position c.55, all three SNVs yielded both aberrant and WT transcripts (Table 1). Notably, the variant with the lowest DL score, c.55G>C (0.34), also had the lowest ratio of aberrant to WT transcripts. At c.56, the c.56G>A variant, with no scores above 0.20, produced only WT transcripts. In contrast, c.56G>C and c.56G>T, with DL scores of 0.29 and 0.46 respectively, yielded both transcript types, and their aberrant/WT transcript ratios aligned with their DL scores. Regarding c.86, c.86A>C, which had the lowest DL score (0.23), did not produce aberrant transcripts. Conversely, c.86A>G, with the highest DL score (0.67), resulted in a significantly higher aberrant/WT transcript ratio of 4.13/1. Interestingly, c.86A>T, with a lower DL score (0.57) than c.86A>G, led to a much lower aberrant/WT transcript ratio (1/5.31) in comparison.

We then hypothesized that conducting a comprehensive cross-comparison of various events within the same exon context might yield valuable insights into the remaining intermediate Δ scores. We explored this hypothesis within the contexts of the four exons.

#### Exon 1

All three SNVs at the last nucleotide of exon 1, c.55, exhibited DL scores ranging from 0.34 to 0.51 and DG scores between 0.33 and 0.37 (Table 1). Based on their corresponding mRNA positions, these SNVs were predicted to disrupt the physiological GT splice donor site at positions c.55+1_2 and activate an upstream cryptic splice donor site within exon 1 (i.e., the GT dinucleotide at position c.7_8) (Fig. 3a). This would result in a significantly shortened transcript that lacked the last 49 nucleotides (i.e., c.7 to c.55) of exon 1. Interestingly, our FLGSA assay detected an aberrant transcript that retained the first 140 bases of intron 1 (Fig. 2), due to the activation of a downstream cryptic GT splice site located at the deep intron 1 region (precisely at c.55+141_142) (Fig. 3a).

**Figure 3.**
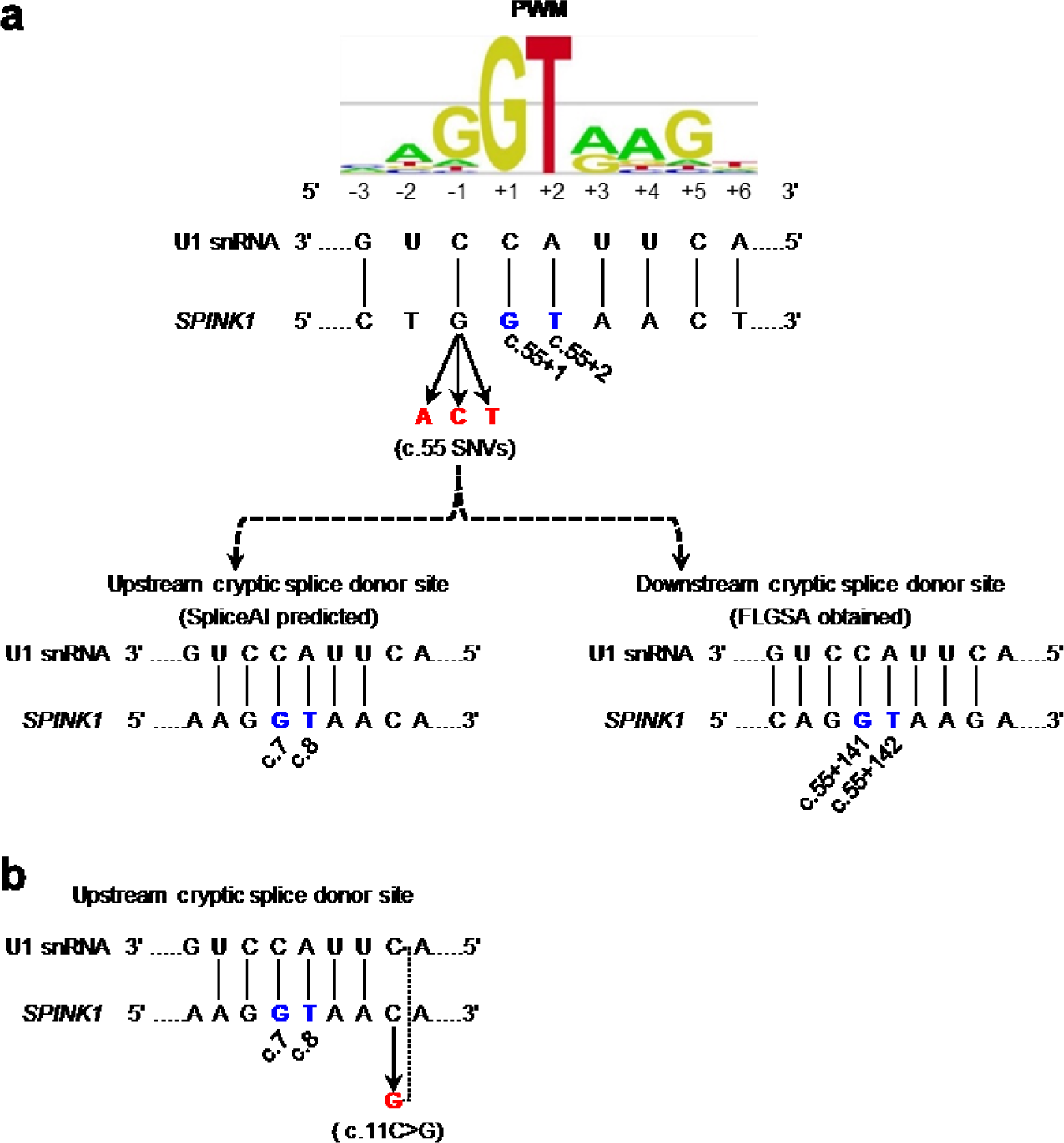
Interpretation of the three c.55 SNVs and the c.11C>G variant in exon 1 by reference to SpliceAI predictions and FLGSA results. **a** Illustration of the (partial) disruption of the physiological 5’ splice donor site of *SPINK1* intron 1 caused by the three potential SNVs at the last nucleotide of exon 1 (c.55). This disruption is shown in the context of the corresponding 9-bp 5’ splice signal sequence, which interacts with the 3’-GUCCAUUCA-5’ sequence at the 5’ end of U1snRNA. SpliceAI predicted this disruption (DL scores, 0.34 to 0.51) and the activation of an upstream cryptic splice donor site within exon 1 (DG scores, 0.33 and 0.37). However, our FLGSA assay revealed the activation of a downstream cryptic splice donor site. Vertical lines indicate paired bases between the 9-bp 5’ splice signal sequence and the 5’ end sequence of U1snRNA. The GT dinucleotides involved are highlighted in blue, with their positions (in accordance with NM_001379610.1) indicated. The 9-bp 5’ splice site signal sequence position weight matrices (PWM) were sourced from Leman et al. [44], an Open Access article distributed under the terms of the Creative Commons Attribution Non-Commercial License. Note that the 9-bp 5’ splice signal sequences, whether in the context of the consensus sequence or *SPINK1* sequences, are presented in DNA. **b** Illustration of the c.11C>G variant in the context of the aforementioned upstream cryptic splice donor site. A dotted line represents the new base pairing derived from the variant, enhancing the interaction between the 9-bp 5’ splice signal sequence and the 5’ end sequence of U1snRNA.

We performed two additional analyses to address the discrepancy between *in silico* predictions and experimental data. First, considering that the Illumina precomputed scores were created using a maximum distance of 50 bp, we reevaluated the three possible c.55 SNVs using SpliceAI with an extended distance of 10,000 bp and the hg38 sequence [40]. Although the resulting DG and DL scores showed slight variations compared to the Illumina precomputed scores (Supplementary Table S1), they did not alter the predicted splicing outcomes. Second, we examined the predicted and experimentally obtained cryptic GT donor site in the context of the 9-bp 5’ splice signal sequence and their pairing with the 3’-GUCCAUUCA-5’ sequence at the 5’ end of U1snRNA (see [32] and references therein). Interestingly, the experimentally identified cryptic GT donor site was found within a 9-bp 5’ splice signal sequence that exhibited 8 bp complementarity with the 9 bp U1snRNA sequence. In contrast, the *in silico* predicted cryptic GT donor site resided within a 9-bp 5’ splice signal sequence with only 6 bp complementarity to the 9 bp U1snRNA sequence (Fig. 3a). It’s noteworthy that this *in silico* predicted cryptic GT donor site coincides with the previously discussed false physiological GT dinucleotide at position c.7_8, which was related to the DL score of the known c.26T>G variant (see *Retrospective correlation of FLGSA data with SpliceAI predictions for known SPINK1 coding SNVs*). Based on these new findings, we speculate that SpliceAI might have favored the nearby cryptic donor site in exon 1 over the more distant cryptic donor site in intron 1.

Another variant in exon 1, c.11C>G, was predicted to induce a splicing effect similar to the three potential c.55 SNVs based on the SpliceAI scores. Interestingly, it displayed even higher DG and DL scores than those of the three potential c.55 SNVs (Table 1). However, the FLGSA analysis did not reveal aberrant transcripts associated with the c.11C>G variant. As illustrated in Fig. 3b, c.11C>G resides within the 9-bp 5’ splice signal sequence linked to the previously mentioned cryptic 5’ splice GT donor site at positions c.7_8. Notably, it increased sequence complementarity with the 9-bp U1snRNA sequence from 6 bp to 7 bp compared to the WT sequence. It’s essential to highlight that, in this scenario, the physiological intron 1 splice donor signal sequence remains unaltered. Bearing this in mind, we conjecture that the enhanced sequence complementarity brought about by the c.11C>G variant may have encountered difficulties in competing with the intact physiological intron 1 splice donor signal sequence, which exhibited 8 bp complementarity with U1snRNA (Fig. 3a).

Shifting our focus to other variants, let’s consider c.15C>T, which had the highest DG score (0.52) among all possible coding SNVs in exon 1, but it only had a DL score of 0.13. We also have c.43T>G, which had a DG score of 0.38 but a DL score of zero. Importantly, neither of these variants resulted in the generation of aberrant transcripts (Table 1). Drawing parallels with the previously discussed c.11C>G variant, we propose that their predicted cryptic 5’ splice donor sites, located within the coding sequence of exon 1, may not have effectively competed against the intact physiological intron 1 splice donor signal sequence.

#### Exons 2-4

Moving on to exons 2-4, two variants warrant closer examination: c.80G>T in exon 2 and c.178T>G in exon 3. The former variant displayed a DG score of 0.61 but a DL score of only 0.10, while the latter had a DG score of 0.26 but a DL score of zero (Table 1). Our FLGSA analysis did not produce any aberrant transcripts associated with either of these variants. In line with our earlier observations concerning variants in exon 1, such as c.11C>G and c.15C>T, we propose that the predicted cryptic 5’ splice donor sites may not have effectively competed with the intact physiological intron 2 and intron 3 splice donor signal sequences, respectively.

### Extrapolation to unanalyzed *SPINK1* coding SNVs

Finally, we addressed a critical question: Can we reasonably interpret the potential splicing effects of the 658 *SPINK1* coding variants that have not yet undergone functional analysis, based on insights derived from the cross-correlation of FLGSA data and SpliceAI predictions of the 27 known and 35 newly analyzed SNVs? To accurately answer this question, we initially evaluated whether the Illumina precomputed scores significantly deviated from those calculated using a distance of 10,000 bp and the hg38 sequence. Consequently, we manually acquired these latter scores from [40] for all SNVs that underwent FLGSA. While we did observe slight disparities between the two datasets for many SNVs (see Supplementary Table S1), it’s crucial to emphasize that these variations were not expected to result in any changes to the predicted splicing outcomes. Thus, we proceeded confidently, utilizing the Illumina precomputed scores for our subsequent discussions within the contexts of the four exons. Our primary focus remained on unanalyzed SNVs with a Δ score falling within the range of 0.20 to 0.30. This choice was motivated by two factors: (i) we have already included all variants with a physiologically relevant Δ score exceeding 0.30 for FLGSA analysis, and (ii) a Δ score below 0.20 is highly unlikely to impact splicing.

#### Exon 1

In exon 1, we identified nine unanalyzed SNVs with a physiologically plausible Δ score of ≥0.20 (c.3G>A, c.4A>C, c.11C>T, c.14G>T, c.28A>T, c.29G>T, c.36G>A, c.37G>T, and c.45G>T). Notably, these scores consistently fall within the DG type, ranging from 0.20 to 0.24. Interestingly, all of these variants were predicted to have cryptic GT splice donor sites that coincide with the previously discussed GT at c.7_8. Additionally, these variants exhibited low DL scores, spanning from 0 to 0.12. When comparing these scores with those of the functionally analyzed exon 1 SNVs (see Table 1), we can conclude that none of these nine variants had any discernible impact on splicing.

#### Exon 2

In exon 2, we identified only two unanalyzed SNVs with a physiologically plausible Δ score of ≥0.20 (c.64G>T and c.81A>T). Both scores are identical (0.22) and belong to the AL type. AG and DG scores of the two variants are zero, while their DL scores are similar (0.11-0.13). Evaluation of the corresponding mRNA positions associated with the AL and DL scores demonstrated that their predicted splicing outcomes would result in exon 2 skipping.

Of the functionally analyzed exon 2 variants, c.85G>T most closely resembles c.64G>T and c.81A>T in terms of the Δ scores. However, c.85G>T had both slightly higher AL and DL scores than c.64G>T and c.81A>T (AL, 0.25 vs. 0.22; DL, 0.17 vs. 0.11-0.13) and produced no aberrant transcripts. c.65G>T is the next variant that most closely resembles c.64G>T and c.81A>T. c.65G>T had a higher AL score (0.31) but equal DL score (0.17) compared to c.85G>T and generated aberrant transcripts. However, the aberrant transcript was generated alongside the WT transcript, and its amount was much less than that of the WT transcript (ratio of 1:5.16).

Based on this cross-comparison, we can conclude that c.64G>T and c.81A>T are highly unlikely to generate aberrant transcripts.

#### Exons 3 and 4

In exon 3 and 4, none of the functionally analyzed SNVs generated aberrant transcripts. Moreover, except for c.92A>G in exon 3, which had an AG score of 0.13, none of the unanalyzed SNVs had a SpliceAI score exceeding 0.05. Consequently, all SNVs in these two exons were considered not to impact splicing.

### Further validation

While we had confidence in our above extrapolation, we opted for additional validation. Therefore, we selected five variants with the highest scores among those not previously functionally analyzed within exons 1, 2, and 3 for FLGSA analysis. Specifically, they included two of the nine exon 1 variants mentioned above (c.29G>T and c.37G>T), the two exon 2 variants mentioned earlier (c.64G>T and c.81A>T), and the exon 3 variant c.92A>G (Table 2). As shown in Fig. 4, all five variants exclusively produced WT transcripts, thereby validating our extrapolation.

**Figure 4.**
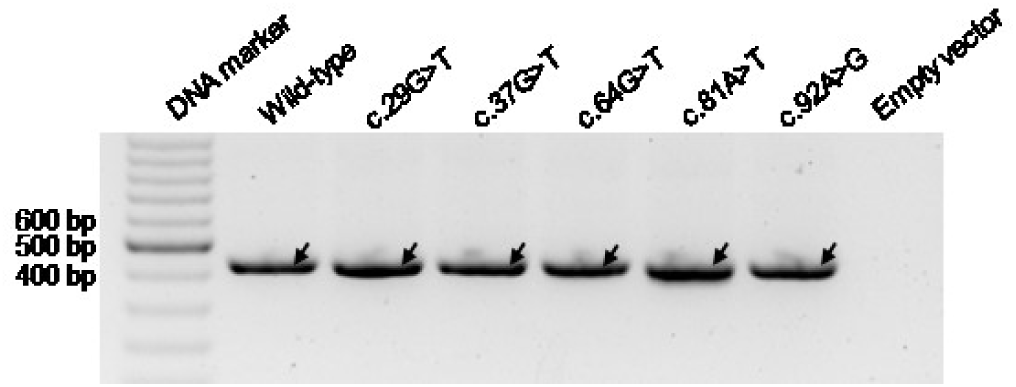
RT-PCR results for the validation analysis of five potential *SPINK1* coding variants through FLGSA. Arrows indicate wild-type transcripts, all of which were confirmed by Sanger sequencing. FLGSA, full-length gene splicing assay. RT-PCR, reverse transcription-PCR.

**Table 2.**
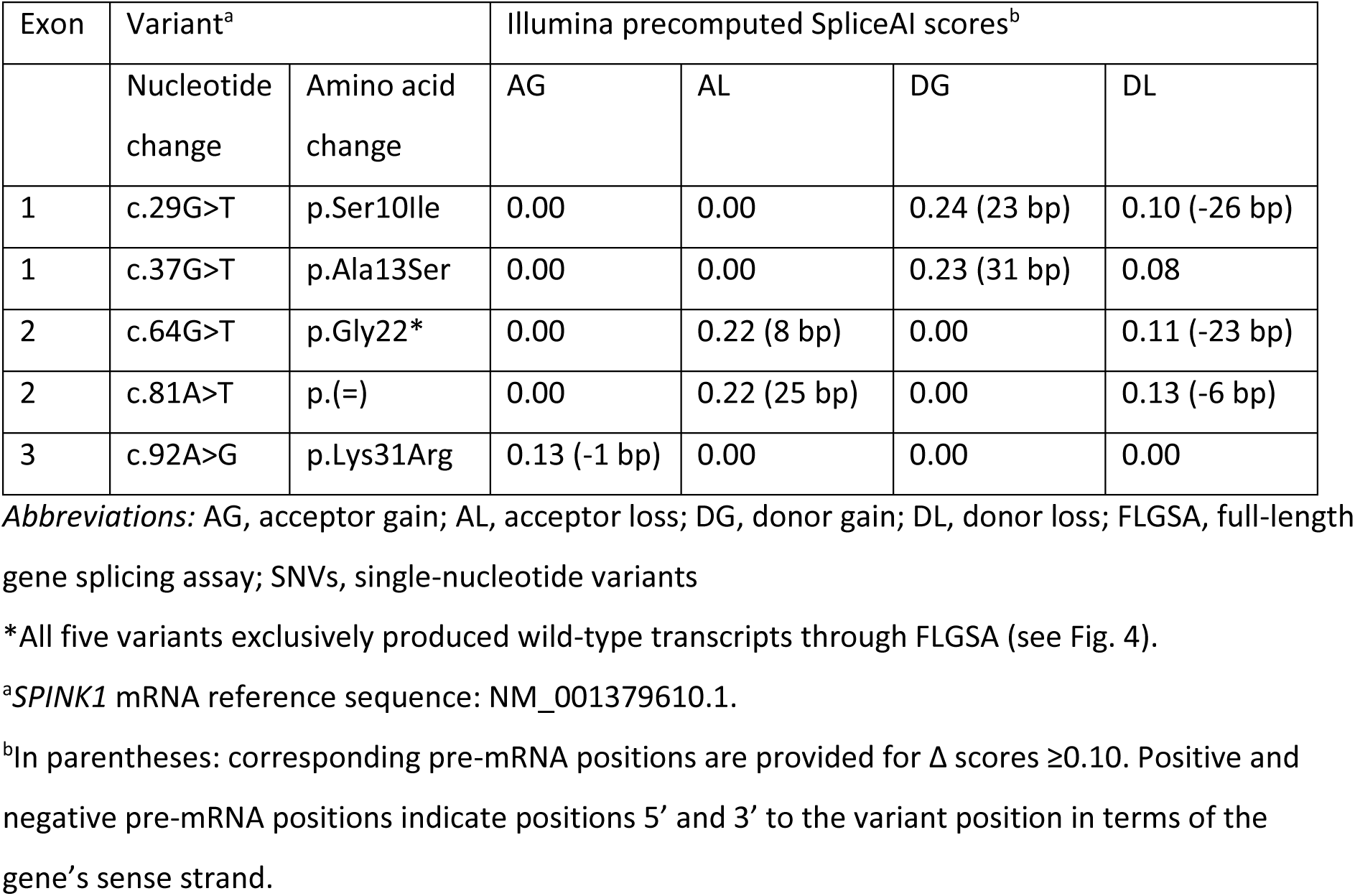
Selected five *SPINK1* coding SNVs for further validation*.

### Overview of splice-altering coding SNVs in the *SPINK1* gene

Overall, our study covers 67 *SPINK1* coding SNVs, accounting for 9.3% of all 720 possible coding SNVs and affecting 46 (19.2%) of 240 coding nucleotides. Out of these 67 SNVs, 12 were experimentally found to impact splicing. Based on a comprehensive cross-correlation of FLGSA-obtained and SpliceAI-predicted data, we conclude that all unanalyzed potential coding SNVs in the *SPINK1* gene are unlikely to have a significant effect on splicing. Therefore, the 12 splice-altering events identified in our study represent the totality of splice-altering events among the 720 potential coding SNVs in the *SPINK1* gene.

Among the 12 splice-altering events, nine produced both WT and aberrant transcripts, while the remaining three exclusively generated aberrant transcripts. These splice-altering SNVs were predominantly found in exons 1 and 2, particularly affecting the first and/or last coding nucleotide of each exon. Among the splice-altering events, 11 were missense variants, accounting for 2.17% of the 506 potential missense variants, while one was synonymous, accounting for 0.61% of the 164 potential synonymous variants (see Table 3).

**Table 3.**
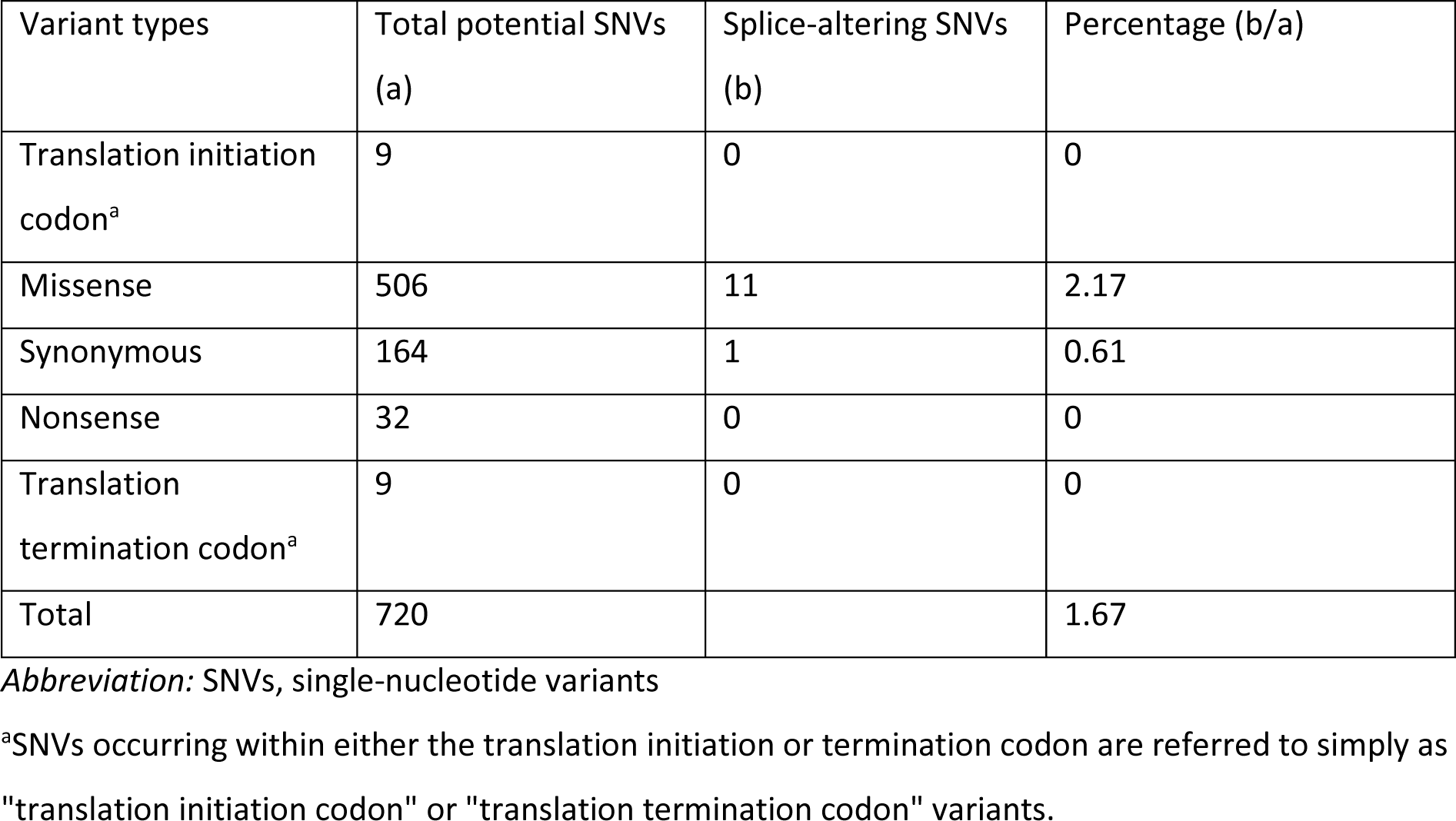
Summary of splice-altering coding SNVs in the *SPINK1* gene.

## Discussion

In this study, we leveraged the well-established FLGSA assay [7, 27–31] in conjunction with SpliceAI [11] to explore the prospective interpretation of splicing effects for all potential coding SNVs within the *SPINK1* gene, following the structured progression outlined in Fig.1.

Our analysis unveiled intriguing discrepancies between SpliceAI predictions and the FLGSA data. Notably, SpliceAI erroneously predicted that several exon 1 SNVs would disrupt a splice donor site within the coding sequence of exon 1 (see Table 1). These predictions conflicted with two critical pieces of evidence: Firstly, only one known *SPINK1* transcript isoform, NM_001379610.1, is expressed in the exocrine pancreas [22, 23]. Secondly, none of the other four alternative *SPINK1* transcript isoforms [43] were found to utilize a GT dinucleotide within the exon 1 coding sequence as a splice donor site. The convergence of these findings, along with the results of our FLGSA assay, strongly suggests that these SpliceAI predictions were indeed spurious.

Another significant discrepancy centered around the splicing outcomes of the three SNVs at c.55, specifically involving the last nucleotide of exon 1. SpliceAI predicted the activation of an upstream cryptic GT dinucleotide at c.7_8, whereas our FLGSA assay identified the activation of a downstream cryptic GT dinucleotide located at c.55+141_142. Intriguingly, the SpliceAI-predicted cryptic GT dinucleotide coincided with one of the aforementioned erroneous splice GT donor sites, highlighting a recurring issue with SpliceAI predictions in the context of exon 1 coding sequences. Notably, our experimentally identified cryptic GT dinucleotide exhibited stronger complementarity with the 3’-GUCCAUUCA-5’ sequence at the 5’ end of U1snRNA, featuring eight complementary bases, in contrast to the SpliceAI-predicted cryptic GT dinucleotide with only six complementary bases (refer to Fig. 3a). It’s important to mention that our experimentally identified cryptic GT dinucleotide is situated more distantly (141 bp) from c.55 than the SpliceAI-predicted cryptic GT dinucleotide (47 bp), potentially explaining why the latter was not correctly predicted by SpliceAI. Additionally, it’s worth acknowledging that variants in exon 1 are not readily amenable to analysis through the commonly used minigene assay [8], and the activation of cryptic donor or splice sites in deep intronic regions may often elude detection via a minigene assay.

Except for the aforementioned discrepancies, we found a robust correlation between FLGSA data and SpliceAI predictions, particularly concerning the 0.20 and 0.80 threshold scores and the findings for all possible SNVs at c.55, c.56, and c.86. While the relationship between FLGSA data and SpliceAI scores below 0.20 or above 0.80 tended to be straightforward, deciphering the correlation between FLGSA data and intermediate SpliceAI scores within the range of 0.20 to 0.80 presented challenges. Nevertheless, our efforts to correlate these intermediate SpliceAI scores with FLGSA findings yielded intriguing insights. For instance, variants with intermediate SpliceAI scores, when leading to aberrant transcripts, often produced a mix of aberrant and WT transcripts. Furthermore, in the case of all possible SNVs at c.55, c.56, and c.86, intermediate SpliceAI scores seemed to correlate with the aberrant/WT transcript ratio. These mutually reinforcing data were instrumental in guiding a cross-comparison concerning unanalyzed variants, allowing us to reasonably extrapolate that none of the unanalyzed coding SNVs in the *SPINK1* gene are likely to exert a significant effect on splicing.

To the best of our knowledge, this study represents the first attempt to prospectively interpret all potential coding SNVs in a disease-associated gene. Our findings unveiled that within the *SPINK1* gene, 2.17% of all potential missense variants, 0.61% of all potential synonymous variants, but none of the potential nonsense variants have an impact on splicing. In total, 1.67% (12 out of 720) of all potential coding SNVs in the *SPINK1* gene were found to alter splicing.

Among the 12 splice-altering variants, five (c.84A>G, c.86A>G, c.87G>A, c.87G>C, and c.87G>T) led exclusively or predominantly to aberrant transcripts. These five variants can be classified as “pathogenic” and would have been mislabeled as silent or missense variants without the FLGSA assay. The remaining seven variants exhibited aberrant to WT transcript ratios ranging from 1/21.72 to 2.97/1. In all these cases, the aberrant transcripts—either retaining part of intron 1 or omitting the entire exon 2—would yield a non-functional product. However, when the aberrant transcript ratio is substantially lower than that of the WT transcript, the variant in question (e.g., c.55G>C with a ratio of 1/21.72) may not be of pathogenic significance.

It’s important to acknowledge the limitations of our FLGSA assay. For instance, like the minigene assay, our experiments required transfected cells, which may not always faithfully recapitulate *in vivo* conditions. However, findings from correlation of our FLGSA data with SpliceAI and cross-comparisons of our FLGSA data across different variants gave strong support to the validity of our FLGSA assay.

## Conclusions

By integrating the FLGSA assay with SpliceAI predictions, our study presents compelling evidence that 1.67% of potential *SPINK1* coding SNVs exert a discernible impact on splicing outcomes. Our findings underscore the critical necessity of conducting splicing analysis within the broader genomic context of the target gene, a perspective that can reveal splicing outcomes often missed by conventional minigene assays. Additionally, we emphasize the inherent uncertainties associated with intermediate SpliceAI scores (ranging from 0.20 to 0.80), highlighting the critical role of functional analysis in variant interpretation. Finally, our approach offers potential implications for transitioning from “retrospective” to “prospective” variant analysis in other disease genes, accelerating the full realization of precision medicine in the whole exome sequencing or whole genome sequencing era.

## Supporting information

Supplementary Tabls S1

## Data Availability

All data produced in the present work are contained in the manuscript.

## Funding

This research was funded by the National Natural Science Foundation of China (81800569 to HW, 82000611 to J-HL, and 82000606 to X-YT), the Shanghai Pujiang Program (2020PJD061 to J-HL), the Shanghai Sailing Program (20YF1459400 to X-YT). Support for this study also came from the Institut National de la Santé et de la Recherche Médicale (INSERM), the Association des Pancréatites Chroniques Héréditaires and the Association Gaétan Saleün, France. The funding bodies did not play any role in the study design, collection, analysis, and interpretation of data or the writing of the article and the decision to submit it for publication.

## Authors’ contributions

HW, JHL, XYT, and WBZ designed the study, conducted the experiments, and contributed to paper writing. SS acquired the Illumina precomputed SpliceAI scores. EM, YF, GLC, and CF contributed to the conception of the study. ZL was involved in study conception and coordinated the experiments. JMC conceived the study, obtained the latest SpliceAI scores, and wrote the manuscript. All authors participated in data review, revised the manuscript with critical intellectual input, approved the submitted version, and agreed to be personally accountable for the author’s own contributions and to ensure that questions related to the accuracy or integrity of any part of the work, even ones in which the author was not personally involved, are appropriately investigated, resolved, and the resolution documented in the literature.

## Acknowledgements

The authors are grateful to GENEWIZ Biotech Co. (Suzhou, China) for their assistance in preparing the variant expression vectors.

## Notes

### Competing Interest Statement

The authors have declared no competing interest.

